# The effectiveness of the first dose of BNT162b2 vaccine in reducing SARS-CoV-2 infection 13-24 days after immunization: real-world evidence

**DOI:** 10.1101/2021.01.27.21250612

**Authors:** Gabriel Chodick, Lilac Tene, Tal Patalon, Sivan Gazit, Amir Ben Tov, Dani Cohen, Khitam Muhsen

## Abstract

**Background:** BNT162b2 vaccines showed high efficacy against COVID-19 in a randomised controlled phase-III trial. A vaccine effectiveness evaluation in real life settings is urgently needed, especially given the global disease surge. Hence, we assessed the short-term effectiveness of the first dose of BNT162b2-vaccine against SARS-CoV-2 infection. Given the BNT162b2 Phase-III results, we hypothesized that the cumulative incidence of SARS-CoV-2 infection among vaccinees will decline after 12 days following immunization compared to the incidence during the preceding days.

**Methods:** We conducted a retrospective cohort study using data from 2·6 million-member state-mandated health provider in Israel. Study population consisted of all members aged 16 or above years who were vaccinated with BNT162b2-vaccine between December/19/2020 and January/15/2021. We collected information regarding medical history and positive SARS-CoV-2 polymerase chain reaction test from days after first dose to January/17/2021. Daily and cumulative infection rates in days 13-24 were compared to days 1-12 after first dose using Kaplan-Meier survival analysis and generalized linear models.

**Findings:** Data of 503,875 individuals (mean age 59·7 years SD=14·7, 47·8% males) were analysed, of whom 351,897 had 13-24 days of follow-up. The cumulative incidence of SARS-CoV-2 infection was 0·57% (n=2_484_) during days 1-12 and 0·27% (n=614) in days 13-24. A 51·4% relative risk reduction (RRR) was calculated in weighted-average daily incidence of SARS-CoV-2 infection from 43·41-per-100,000(SE=12·07) in days 1-12 to 21·08-per-100,000(SE=6·16) in days 13-24 following immunization. The decrement in incidence was evident from day 18 after first dose. Similar RRRs were calculated in individuals aged 60 or above (44.5%), younger individuals (50.2%), females (50.0%) and males (52.1%). Findings were similar in sub-populations and patients with various comorbidities.

**Conclusions:** We demonstrated an effectiveness of 51% of BNT162b2 vaccine against SARS-CoV-2 infection 13-24 days after immunization with the first dose. Immunization with the second dose should be continued to attain the anticipated protection.

**Research in context:** *Evidence before this study:* We searched PubMed for follow-up studies regarding the effectiveness of BNT162b2 mRNA Covid-19 Vaccine without any language restrictions. The search terms were (BNT162b2 OR mRNA Covid-19 Vaccine) AND (effectiveness OR real-world OR phase IV) until Jan 15, 2021. We found no relevant observational studies among humans. We also assessed Phase II and Phase III clinical trials with BNT162b2 mRNA vaccine.

*Added value of this study:* To our knowledge, this is the first and largest phase IV study on the effectiveness of the BNT162b2 mRNA COVID-19 vaccine in real-world settings. Our findings showed that the first dose of the vaccine is associated with an approximately 51% reduction in the incidence of PCR-confirmed SARS-CoV-2 infections at 13 to 24 days after immunization compared to the rate during the first 12 days. Similar levels of effectiveness were found across age groups, sex, as well as among individuals residing in Arab or ultra-orthodox Jewish communities that display an increased COVID-19 risk.

*Implications of all the available evidence:* The study results indicate that in real life the first dose of the new BNT162b2 mRNA COVID-19 vaccine confers around 50% protection against overall SARS-CoV-2 infections (symptomatic or asymptomatic). Together our findings and the 95% efficacy shown in the phase III trial, suggest that the BNT162b2 vaccine should be administered in two doses to achieve maximum protection and impact in terms of disease burden reduction and possibly reducing SARS-CoV-2 transmission. COVID-19 vaccines should be urgently deployed globally.

## Introduction

The recently authorized BNT162b2 COVID-19 vaccine has demonstrated 95% efficacy in preventing COVID-19 with the two-dose regimen in phase III randomized placebo-controlled trial (RCT), with the second dose given 21 days after the first vaccine dose [1]. The protection of the vaccine against the disease was already evident 12 days after the first dose. The European Medicines Agency (EMA) has approved the BNT162b2 emergency use using two doses separated by at least 21 days[2].

In light of the peaking outbreak of COVID-19, the UK authorities decided to vaccinate a large number of at-risk people in the shortest possible time by postponing the second dose towards the end of the recommended vaccine dosing schedule of 12 weeks[3]. The same immunization approach is also contemplated in other countries and by the World Health Organisation (WHO) in view of limited doses of vaccine available to date for mass vaccination [4]. Accordingly, there is an urgent global need to understand the real-world short-term effectiveness of the vaccine after the first dose.

The BNT162b2 RCT showed vaccine efficacy of 52% (95% Confidence intervals [CI] 29·5 to 68·4) after immunization with the first dose. This is comparable with the minimal acceptable level of efficacy of 50% in preventing COVID-19 as indicated by the WHO [5] and by the US Food and Drug Administration [6] as one of the essential criteria to confer Emergency Use Approval (EUA) to COVID-19 candidate vaccines. However, the effectiveness of the new vaccine in protecting against infection is difficult to assess in phase III clinical trials. Instead, it requires large phase IV studies in real-world settings where the vaccine is widely deployed [5]. Moreover, the lack of systematic testing for SARS-CoV-2 infection in vaccinees and placebo recipients as part of the protocol leaves important gaps in knowledge regarding the vaccine efficacy in preventing asymptomatic infection and consequent viral transmission. Moreover, assessment of the vaccine effectiveness outside clinical trial setting, in real life, is warranted especially given the complex and unusual storage and handling requirements of BNT1622b2 vaccine.

In Israel, vaccination against coronavirus using BNT162b2 mRNA vaccine started on December 19, 2020, with priority given to individuals over 60 years old, healthcare workers, and clinically-vulnerable patients. As of January 15, 2021, Israel ranks first in vaccination doses with 25·3 per 100-capita [7]. The aim of this study is to assess the short-term effectiveness of one dose of BNT162b2 vaccine in reducing infection with SARS-CoV-2 in real-world settings. In light of the RCT results, our hypothesis was that the cumulative incidence of SARS-CoV-2 infection among individuals who received BNT162b2 vaccine will decline after 12 days following immunization compared to the incidence during the preceding 1-12 days.

## Methods

### Study design and data sources

We conducted a retrospective cohort study using data from a single healthcare provider to estimate the short-term effectiveness of the first dose of the BNT162b2 vaccine against SARS-2-CoV-2 infection. Data sources were the central databases of Maccabi Healthcare Services (MHS). MHS is a 2·6 million-member state-mandated, not-for-profit, sick fund in Israel, representing a quarter of the Israeli population. Membership in sick funds is compulsory in Israel, and by the National Health Insurance Law of 1994, all citizens must freely choose one of four national sick funds that are prohibited by law from denying membership to any Israeli resident. The dataset included extensive demographic data, anthropometric measurements, clinic and hospital diagnoses, medication dispensed, and comprehensive laboratory data from a single central laboratory. In Israel, everyone is assigned a unique person-specific alphanumeric identifier called a National Health Index Number, which is used across all health systems, including in these datasets, thereby enabling data linkage.

We used data linkage to assign vaccine exposure, collect information regarding medical history and positive SARS-CoV-2 polymerase chain reaction (PCR) test [8]. PCR tests for SARS-CoV-2 are obtained from nasopharyngeal swabs. PCR testing is offered for all citizens free of charge, mostly without a need for referral.

We obtained ethical approval from Maccabi Healthcare Services Ethics Committee.

### Study population and endpoint

Our study population consisted of all MHS members aged 16 or above years who were vaccinated during a mass immunization program from December 19, 2020, to January 15, 2021 (inclusive). We used as reference the results of the phase III [1] that provide experimental evidence that the BNT162b2 vaccine conferred no or little protection against SARS-CoV-2 infection until day 12 post vaccination with the first dose. Therefore, we calculated cumulative incidence of infection in days 13 to 24 as compared to days one to 12 post vaccination with the first dose. Index day was thus defined as day one after first dose, for days one to 12 cohort and day 13 for days 13 to 24 cohort. Follow-up for infection started from index date and lasted until the date of first positive SARS-CoV-2 PCR result, death, leaving MHS, January 17, 2021, or 12 days after index date. SARS-CoV-2 infection case definition was having at least one record of primary positive SARS-CoV-2 PCR test in the MHS databases. Excluded from analysis were patients who had a documented positive SARS-CoV-2 (n=2,022) prior to index date or individuals who joined MHS after February 2020 and therefore have an incomplete medical history (n=6389).

### Additional variables

Individual-level clinical and demographic data on study cohort were collected from MHS central datasets. This included age at index date, sex, body mass index (BMI) as well as data on underlying diseases from computerized registries including cancer, immunocompromised conditions, diabetes [9], cardiovascular diseases [10], and hypertension [11]. The socioeconomic status (SES) index of member’s enumeration area is based on several parameters including household income, educational qualifications, household crowding, material conditions, and car ownership. Further information on participants’ residential area, including characterization of ultraorthodox Jewish or Arab communities was collected. These variables were selected based on the local epidemiological characteristics of COVID-19 in Israel showing higher incidence in ultraorthodox and Arab communities compared to the general population, and in low vs. high SES communities.

### Statistical analysis

Continuous variables were expressed as means (standards deviations [SD]) and medians (range). Categorical variables were summarized as counts and percentages. Cumulative incidence plots of SARS-Cov-2 infection were created using Kaplan-Meier survival analysis and compared with the log-rank test. The comparison of the incidence of PCR-confirmed SARS-CoV-2 infection between the two study periods, 1-12 and 13-24 days following to immunization with BNT162b2 vaccine was first estimated using a generalized linear models (GLM), applying a negative-binomial distribution with a log-link and log-time-at-risk as an offset. The offset was used to scale the counts of SARS-CoV-2 infections to daily incidence, expressed as cases per 100,000. The dependent variable was the number of SARS-CoV-2 per day during the 12 days of the follow-up for each group. Independent variables were sex and age category. Vaccine effectiveness was defined as infection relative risk reduction (RRR) and calculated as: (1 – relative risk) × 100. Analyses were stratified by age, sex, ultraorthodox Jewish sector/community status, and comorbid condition.

The incidence of COVID-19 in Israel has changed during the study period. Although Israel has been under lockdown during the study period, according to the Israel Ministry of Health data [12] there was a 33·7% increase in the number of laboratory confirmed SARS-CoV-2 cases in adults from week 52/20 to 53/20 and a 35.7% increase between week 53/20 and 01/21. Since the study groups distributed differently along the calendar time, this may have attenuated our estimates of vaccine effectiveness. Therefore, sensitivity analyses included omitting first and last calendar week and censoring at second dose of vaccine. Patient with a positive SARS-CoV-2 test after first dose are recommended to postpone their second dose. In order to avoid a potential selection bias we therefore did not censor the follow-up period at date of second dose. All analyses were done using IBM-SPSS Version 27 (Armonk, NY: IBM Corp) and R packages magrittr, readtext, dplyr, ggplot2, tidyverse, survival, and survminer.

### Role of the funding source

There was no external funding for this analysis.

## Findings

Data of 503,875 individuals (mean age 59·7 years [SD=14·7], 47·8% males) were analysed of whom 351,897 had between 13 to 24 days of follow-up after first dose (Table 1). The total population accounts for 26% of the MHS members aged 16 or above. During the follow-up period 49,814 (9·9%) were tested for SARS-CoV-2.

**Table 1:** Characteristics of study population, by duration of follow-up from day of vaccination with first dose of BNT162b2 COVID-19.

|  | Days from first dose |  |
| --- | --- | --- |
|  | From day 1<br>(n=503,875) | From day 13<br>(n=351,897) |
| **Age** years, mean ± SD | 59.7 ± 14.7 | 61.7 ± 14.4 |
| **Sex** |  |  |
| Males | 239,647 (47.8%) | 170,792 (48.8%) |
| Females | 264,228 (52.4%) | 180,105 (51.2%) |
| **BMI** kg/m<sup>2</sup>, mean ± SD | 27.49 ± 5.00 | 27.38 ± 5.04 |
| **SES\*** [range] | 7 [1-10] | 7 [1-10] |
| **Residential community** |  |  |
| Ultraorthodox Jews | 13,936 (2.8%) | 9,403 (2.7%) |
| Arabs | 13,084 (2.6%) | 7,782 (2.2%) |
| **Comorbidities** |  |  |
| Diabetes mellitus | 86,161 (17.1%) | 67,662 (19.2%) |
| Cardiovascular diseases | 57,057 (11.3%) | 46,235 (13.1%) |
| Hypertension | 186,047 (36.9%) | 144,296 (41.0%) |
| Cancer | 80,496 (16.0%) | 64,861 (18.4%) |
| Immunosuppression | 21,911 (4.3%) | 16,571 (4.7%) |

\* Residential area socioeconomic status on a scale from 1 (lowest) to 10 BMI: Body mass index; SD: standard deviation; SES: socioeconomic status

During the study period a total of 3098 incident cases of PCR-confirmed SARS-CoV-2 infection were identified with a cumulative risk of 0.84% (figure 1), with 0·57% (n=2_484_) occurring during the first follow-up period (days 1-12) and 0·27% (n=614) during the second follow-up period (days 13-24) (Figure 2). The significant decrement in incidence was evident from day 18 after first dose. Using GLM, a RRR of 51·4% (95% CI −7·2% to 78·0%) was calculated with weighted average daily incidence of SARS-CoV-2 infection declining from 43·41 per 100,000 (SE=12·07) in days 1 to 12 to 21·08 (SE=6·16) in days 13 to 24 following immunization with first dose. Similar findings were seen in stratified analyses by age group (RRR=44·5% in individuals aged 60 years or above vs. 50·2% in individuals aged under 60 years), sex (50·0% in females vs. 52·1% in males), and Jewish ultraorthodox communities (53·5%), as well as when first and last calendar weeks were omitted from the analysis (Figure 3).

**Legend to Figure 1:**
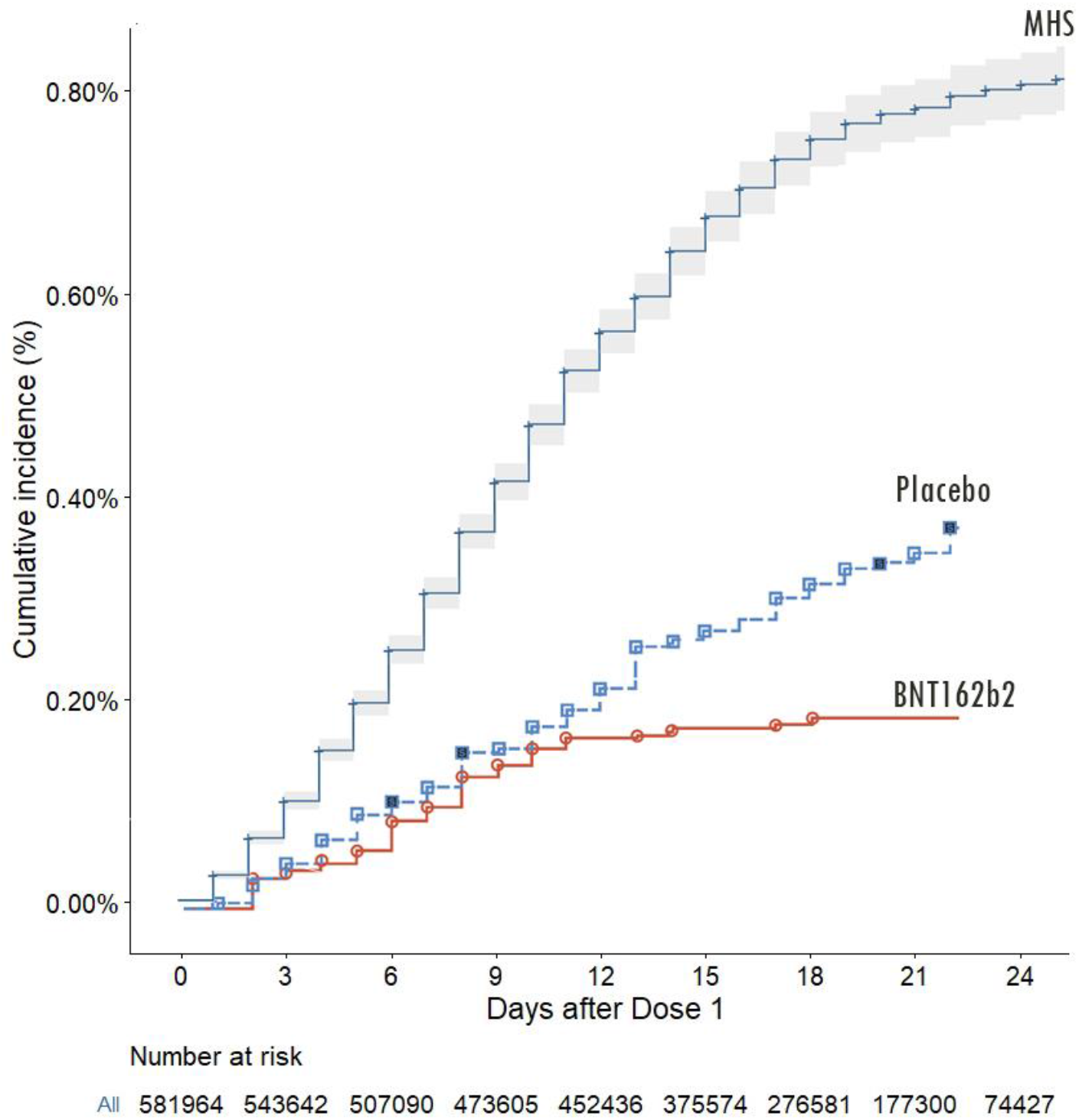
Cumulative incidence of SARS-CoV-2 infection by days since index date in MHS and COVID-19 in BNT162b2 phase III trial (adopted from Polack et al. 2020)

**Legend to Figure 2:**
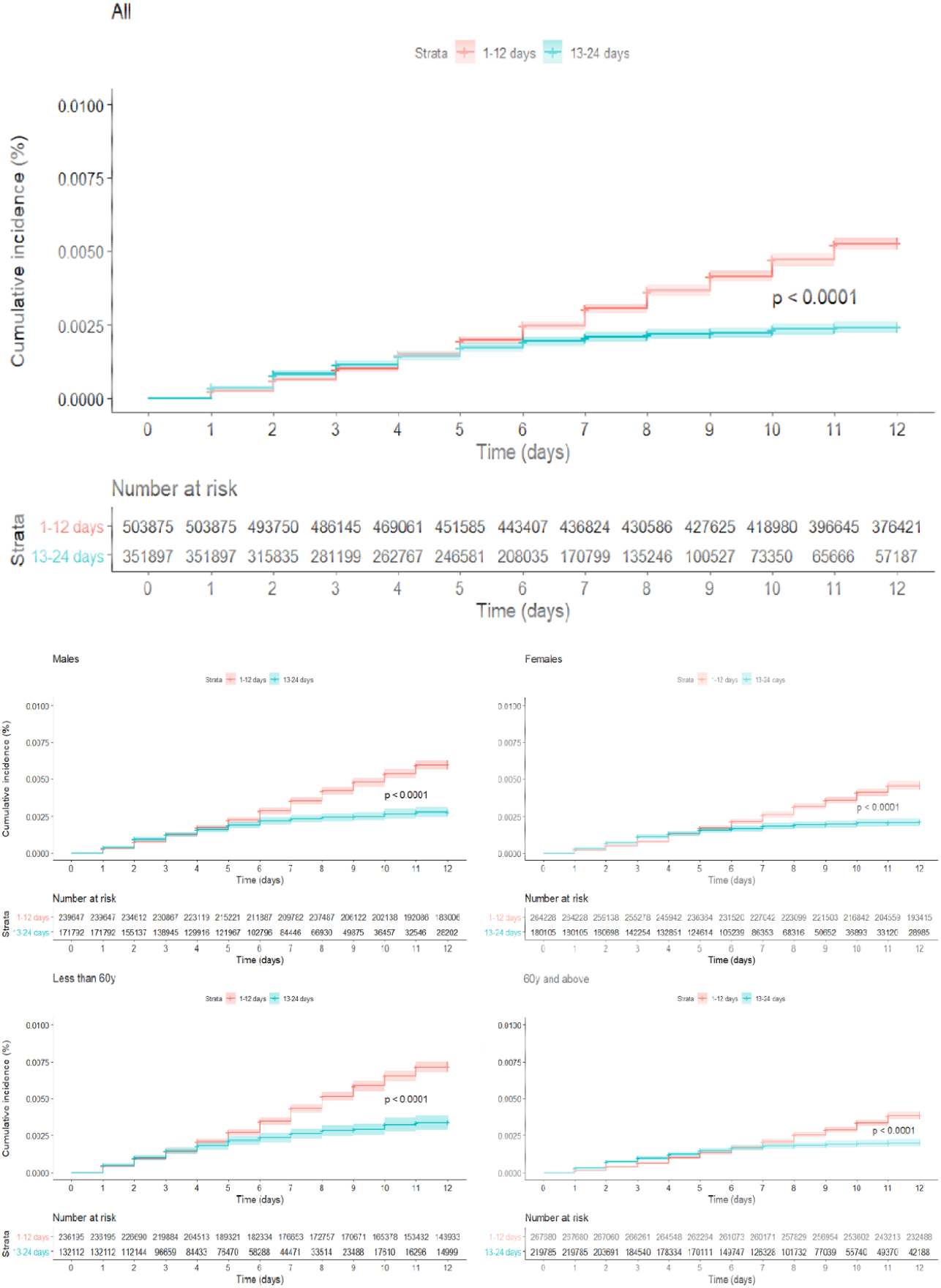
Cumulative incidence of SARS-CoV-2 infection by days since index date in the period of 1 to 12 days after first dose and 13 to 24 days after first dose.

**Legend to Figure 3:**
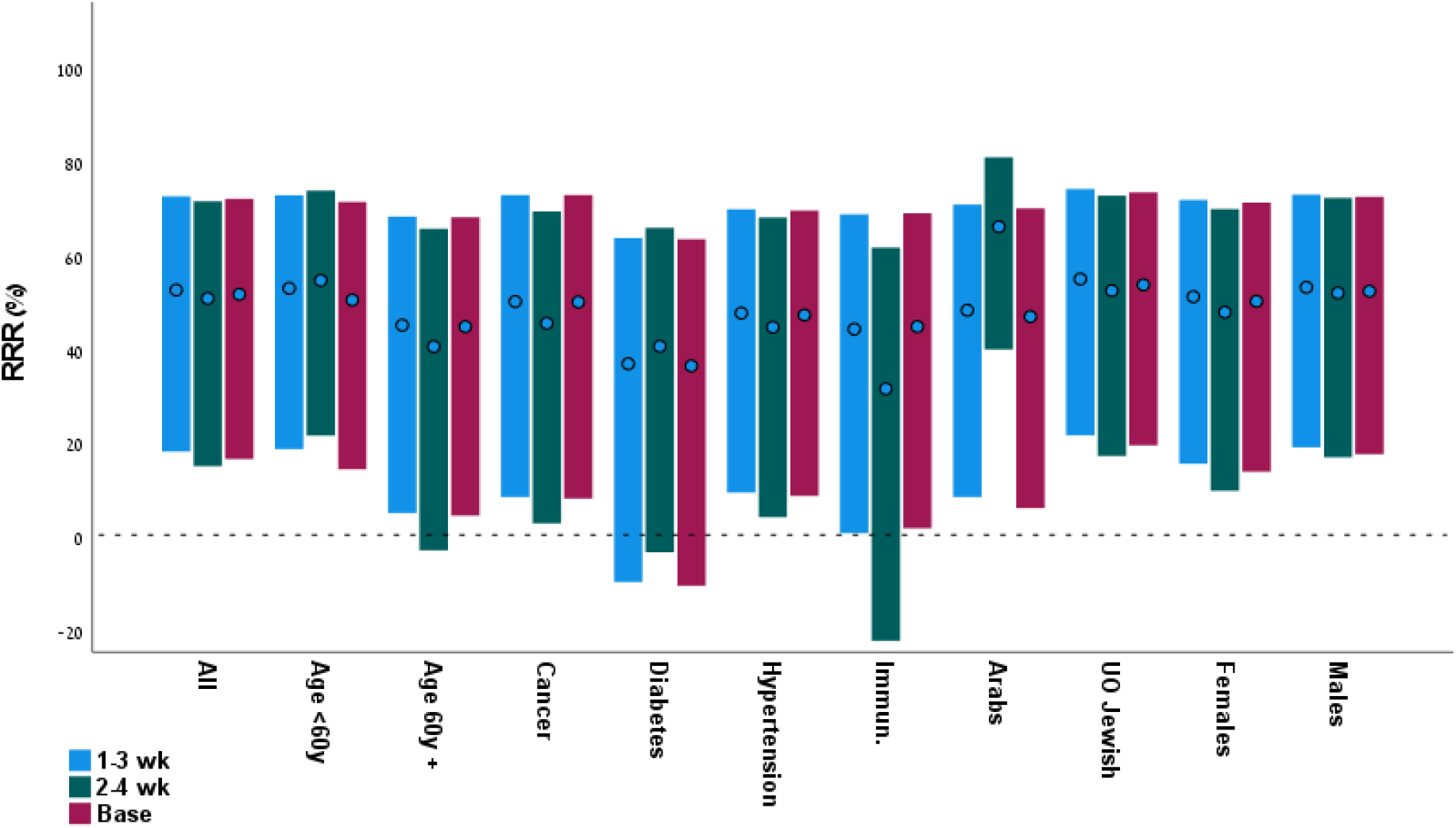
Relative risk reduction and 95%CI by subgroup in base model as well as after excluding first (2-4 weeks) or last (1-3 weeks) calendar week of follow-up period.

## Discussion

In this analysis of real-world data, we demonstrated an effectiveness of approximately 51% of the BNT162b2 mRNA vaccine in reducing the risk of PCR-confirmed SARS-CoV-2 infection 13 to 24 days after immunization with the first dose compared the preceding 1-12 days. While our effectiveness estimate is comparable to the 52% efficacy in preventing COVID-19 calculated in the phase III RCT with BNT162b2[1], there are major differences in interpreting these findings. First, unlike in the RCT, our assessment of vaccine effectiveness relates to days 13 to 24 after first dose whereas the vast majority of cases in the trial occurred earlier. In fact, only two cases were diagnosed between days 14 and 21 days in the vaccinees compared to 18 incident cases in the placebo arm, thus suggesting an efficacy of 89%. Second, our case definition was positive SARS-CoV-2 PCR test irrespective to presenting symptoms that defined COVID-19 cases in the RCT. This might explain the discrepancy in results.

Interestingly, the BNT162b2 vaccine (at 30 mcg, the same dose used in the efficacy study), induced only a modest level of neutralizing antibodies in volunteers aged 18-55 and even lower in volunteers aged 65-85 after the first injection as measured on day 21 of the safety and immunogenicity study [13]. If extrapolated to our estimation of vaccine effectiveness, the immunogenicity data may suggest that these levels of neutralizing antibodies were associated with 90% protective efficacy against COVID-19 and with only around 50% protection against SARS-CoV-2 overall infections 12 to 21 days after immunization with first dose of the BNT162b2 vaccine. Analogically, the very good neutralizing antibody response after the 21 day booster as shown in the immunogenicity study [13] are encouraging suggesting a very good chance for high level protection against SARS-CoV-2 infection as well after immunization with the second dose. It is possible that this very robust systemic functional response 7 days after the second intramuscular injection of BNT162b2 (but not after a single injection) is paralleled by a significant local immune response including both IgG of systemic origin and sIgA, with potential positive implications in prevention of virus transmission. Potent intramuscularly administered conjugate vaccines against bacterial pathogens of the respiratory tract such as *Haemophilus influenzae* type b and *Streptococcus pneumoniae* conjugate were shown to be effective against invasive disease as well as against carriage of serotypes included in the corresponding vaccine formulation[14–17] leading to dramatic direct and indirect protection. Continuing assessment of effectiveness in parallel with serological and virological data in our setting can confirm or refute this assumption and support the evaluation of neutralizing antibodies as correlates of protection against COVID-19 and SARS-CoV-2 asymptomatic infection.

The somewhat lower effectiveness calculated among patients with diabetes is intriguing, particularly in light of a previous reports on impaired immunologic response after seasonal influenza vaccines among patients with diabetes or treated with anti-diabetic medications [18],

Our findings of 51% effectiveness against PCR-confirmed SARS-CoV-2 infection 12 days after immunization with first dose of BNT162b2 in comparison with day 1-12 after vaccine exposure might be an underestimation of the vaccine effectiveness against COVID-19.

Nonetheless, our study provides critically needed evidence on the early performance of BNT162b2 vaccine in real life, and has some important implications in decision making to prevent transmission of SARS-CoV-2 and control the epidemic. While these results are encouraging, the BNT162b2 vaccine should be administered in two dose regimen 21 days apart as licensed for EUA, to achieve maximum protection and impact in reducing COVID-19 devastating burden and possibly the transmission of SARS-CoV-2.

Our study has several strengths. The automated data collection of vaccination status and laboratory results that are offered to all citizens free of charge, allowed us to comprehensively explore vaccine effectives with minimal threat of information bias characterizing studies relying on self-reported diagnosis. By comparing only vaccinated individuals in different time intervals after immunization, we minimized potential selection and indication bias that may stem from comparing vaccinated vs. unvaccinated [19] or test-negative studies [20].

This study also has limitations. In addition, in this database analysis we did not assess COVID-19 symptoms and complications and therefore we were unable to assess effectiveness in protecting against COVID-19. Unreported vaccination is another potential limitation; however, the total number of vaccinated individuals by 1/15/21 in our analysis accounts for approximately 25% [21] of the total number of vaccinated Israelis on 1/15/21 that is comparable to the market share of MHS, leaving little room for significant gap in data.

In conclusion, in this analysis of large-scale real-world data, we showed approximately 51% effectiveness of BNT162b2 COVID-19 vaccine against PCR-confirmed SARS-CoV-2 infection 13-24 days after immunization with the first dose using the preceding 1-12 days as a reference. This estimate was consistent across age, sex, sector, or comorbidity, except for diabetes. These findings have global public health implications and open new horizons towards the control of COVID-19 pandemic and reducing the transmission of SARS-CoV-2. Global effort should accelerate COVID-19 vaccines deployment urgently.

## Data Availability

Data cannot be shared due to regulatory limitations

## Contributors

GC, LT, conceived and designed the study, did the analysis, and took responsibility for the integrity of the data and the accuracy of the data analysis. All authors had full access to all of the data in the study. GC, KM, and DC drafted the manuscript. All authors critically revised the manuscript for important intellectual content and gave final approval for the version to be published. All authors agree to be accountable for all aspects of the work in ensuring that questions related to the accuracy or integrity of any part of the work are appropriately investigated and resolved.

## Declaration of interests

We declare no competing interests.

## Acknowledgments

There was no external funding for this study. We kindly thank Mrs. Esma Herzel and Mr. Hillel Alapi for retrieving the computerized data.

## Notes

### Competing Interest Statement

The authors have declared no competing interest.

### Author Declarations

Maccabi Healthcare Services IRB

